# The predictive value of renal resistance index and plasma cystatin C in pregnancy-related acute kidney injury

**DOI:** 10.1101/2022.10.20.22281289

**Authors:** Suochen Tian, Zhenqin Chang, Qiang Wang, Min Wu, Zhiping Xu, Longying Tian, Yuanda Sui, Yujing Cui, Hui Tian, Xiuli Zou, Mingxin Liu, Tiejun Wu

**Affiliations:** Intensive Care Unit, Liaocheng People’s Hospital, Liaocheng, Shandong, 252000, China; Department of Hemodialysis, Liaocheng People’s Hospital, Liaocheng, Shandong, 252000, China; Department of Healthcare Associated Infection, Liaocheng People’s Hospital, Liaocheng, Shandong, 252000, China; Intelligence Library Center, Liaocheng People’s Hospital, Liaocheng, Shandong, 252000, China

**Keywords:** renal resistance index, cystatin C, creatinine, urine output, pregnancy, acute kidney injury

## Abstract

**Objective:** To analyze the predictive value of renal resistance index (RRI) and plasma cystatin C (pCysC) in pregnancy-related acute kidney injury (PR-AKI).

**Methods:** This study included 182 pregnant women admitted to the intensive care unit (ICU) between May 2016 and June 2021. Intensivists who had received full-time bedside ultrasound Doppler training performed RRI measurements, and blood was drawn to monitor serum creatinine (Scr) and pCysC concentrations. The study continued for 3 consecutive days, marked as the first day, the second day, and the third day, during which the hourly urine output (UO) was monitored and recorded. According to the AKI diagnostic staging criteria, patients with AKI were divided into stages I, II, and III and comprised the study group (Group A), and patients without AKI served as the control group (Group B).

**Results:** Of the 182 enrolled patients, 35 (19.2%) were diagnosed with AKI, including 23 (65.7%) with stage I, 9 with stage II (25.7%), and 3 with stage III (8.6%). Three were excluded owing to the requirement of continuous blood purification. Therefore, 179 patients, 32 in Group A and 147 in Group B, were included. The Scr, pCysC, and RRI of Group A increased on the first, second, and third days, but there was a gradual decrease over time. Each period was compared with the corresponding period in Group B, and there were significant differences (P<0.05). All patients in Group A met the diagnostic criterion of Scr concentration in AKI, and only 34.4% of the patients met the diagnostic criterion of UO. According to the D1 monitoring results, the proportions of increased pCysC and RRI in Group A were 87.5% and 81.3%, respectively. They were significantly different from those in Group B (P<0.001). The three variables of pCysC, RRI, and the combination of pCysC and RRI all independently correlated with AKI. The sensitivity and specificity of pCysC concentration for the prediction of PR-AKI were 87.5% and 84.35%, respectively, and those of RRI were 81.25% and 76.87%, respectively. The sensitivity and specificity of the combination of the two were 96.88% and 72.11%, respectively. Receiver operating characteristic curve analysis showed that these indicators had a significant predictive power for PR-AKI. Although the length of stay in the ICU and hospital in Group A was longer (P<0.05), there was no difference in hospital mortality between the two groups (P>0.05).

**Conclusion:** The diagnosis of PR-AKI based only on Scr and UO was insufficient. RRI and pCysC were important supplements for diagnosing PR-AKI, with good sensitivity and specificity. However, combining the two was better.

## Background

Pregnancy-related acute kidney injury (PR-AKI) is a serious health problem. Effective prevention and early identification of PR-AKI are important, particularly in developing countries [1-4]. In 2007, the Acute Kidney Injury Network Working Group issued a new AKI consensus to better describe and intervene in AKI cases [5]. The diagnostic criterion for AKI was established as an abrupt (within 48 h) reduction in kidney function currently defined as an absolute increase in serum creatinine (Scr) of either ≥ 0.3 mg/dL (≥ 26.4 μmol/L) or a percent increase of ≥ 50% (1.5-fold from that at baseline), or a reduction in urine output (UO) (documented oliguria of < 0.5 mL/kg/h for > 6 h). AKI is divided into stages I, II, and III according to Scr or UO. Since establishing the AKI diagnostic staging standard, many relevant studies have received mixed praises and criticisms. The standard was praised because it further emphasized AKI and its dynamic nature, and it is conducive to early detection and intervention; it was criticized because, in terms of its sensitivity and specificity, it may not be accurate for AKI in different diseases and ethnic groups [6-11]. For patients with severe obstetric conditions admitted to the intensive care unit (ICU), the insufficiency of Scr and UO is more obvious owing to the influence of pregnancy, hypertension, diuretics, and other factors.

According to the current Chinese data [12], the incidence of PR-AKI is 0.02–1.84%. Hypertension during pregnancy (49.2%) and postpartum hemorrhage (13.8%) were the most common causes of PR-AKI. Although the incidence was not high, the population base was large, and special attention is nonetheless required. In another PR-AKI study in a developing country, the incidence of late pregnancy-related renal failure reached 1.78%; pre-eclampsia was the most important cause, followed by postpartum sepsis and stillbirth [13]. There remain concerns in the clinical practice of PR-AKI; nonetheless, an early diagnosis of PR-AKI is important [14].

The renal resistance index (RRI) is monitored using ultrasound Doppler, and plasma cystatin C (pCysC) is a good indicator for predicting AKI. The use of bedside ultrasound for critically ill patients enables quick and timely measurements of RRI. The value of RRI in assessing the risk and prognosis of AKI in critical cases has gradually increased [15-17]. Because of the particularity of the pCysC metabolism, many studies have recommended its use in diagnosing AKI in intensive care and emergency medicine; however, there is no consensus [18-25].

In the clinical evaluation of PR-AKI in severely ill women, the correlation among RRI, pCysC, Scr, and UO remains unclear. Therefore, this study aimed to analyze the predictive value of RRI and pCysC in PR-AKI.

## Methods

### Brief flow chart of the study

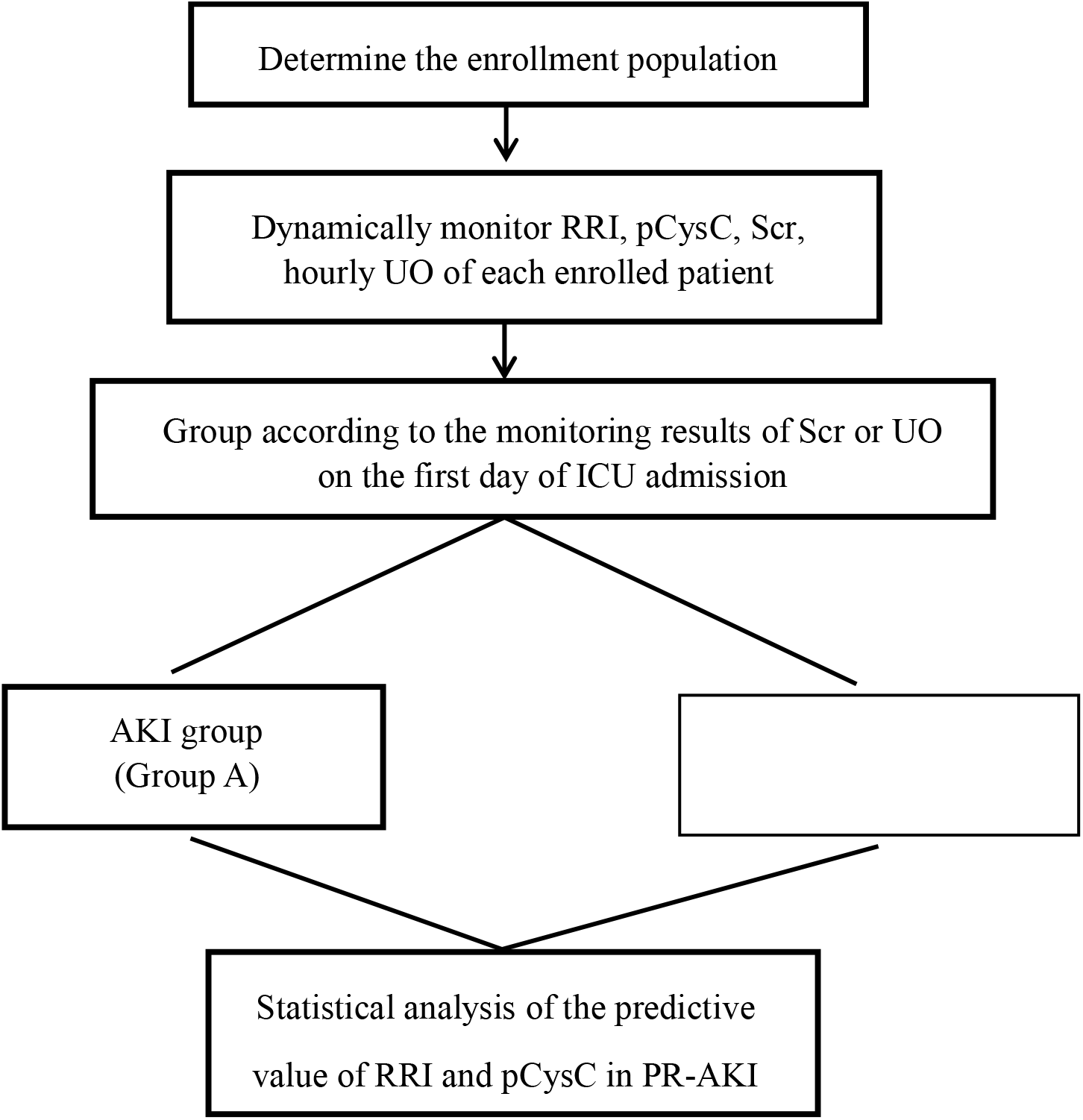

### Study population

The inclusion criteria were (1) aged >18 years, (2) terminated pregnancy, (3) ICU stay > 72 h, and (4) provided signed informed consent. The exclusion criteria were (1) history of chronic renal impairment, congenital heart disease, diabetes, hypertension, pulmonary hypertension, rheumatism, and other chronic diseases; (2) cardiopulmonary resuscitation; (3) rhabdomyolysis; and (4) refusal to participate.

### RRI measurement

First, two intensivists who participated in the study were trained to determine RRI using bedside ultrasound. The instruments used were M8 Super by the Mindray Company (Shenzhen, China). Within 12 h of admission to the ICU, breathing and circulation were relatively stable after the initial treatment. Patients were placed in the supine position, and a convex array probe was used to perform a two-dimensional ultrasound examination. Color Doppler was used in selecting the arch artery or the interlobular artery to determine the peak systolic flow velocity and the minimum diastolic flow velocity from the posterior side of the abdomen. The RRI was calculated according to the following formula: RRI = (peak systolic flow velocity - minimum diastolic flow velocity)/ peak systolic flow velocity. The average RRI of the left and right kidneys was taken as the RRI of each patient. (reference range, 0.58–0.64; upper limit: 0.70). Before the start of the study, 20 ICU patients were randomly selected. Two intensivists performed RRI measurements on these patients. The results of the two measurements were compared and analyzed using SPSS version 23.0. The intra-class correlation coefficient (ICC) was used to evaluate the inter-rater reliability. The ICC of this study was 0.980 [95% CL (0.951–0.992)], and the agreement between the two raters was excellent. The two intensivists thus determined the RRI for the remaining patients who participated in the study.

### Determination of pCysC and Scr

While measuring the RRI, 3 mL of venous blood was drawn to determine the concentrations of pCysC and Scr. The immunoprojection turbidimetric method determined the pCysC concentration (reference range, 0–1.03 mg/L). Scr concentration was determined using the creatinine method (reference range for women, 70–106 µmol/L). The instrument used was the BS400 automatic biochemical analyzer produced by the Mindray Company, and supporting reagents were used.

### UO monitoring

The disposable son–mother type of urine collection bag was used: model specification, 3,100 mL; measuring bottle volume, 500 mL; product code, SK125; urine bag volume, 2600 ml. The responsible nurse entered the hourly UO monitoring results into the ICU information system.

### General principles of monitoring and treatment

All patients will be subjected to dynamic, rapid point-of-care testing (POCT) immediately after being transferred to the ICU. Routine blood biochemical tests will be performed. Acute physiology and chronic health evaluation II (APACHE II) scores will be determined within 24 h. The function of each organ will be dynamically monitored, and the corresponding adjustment or organ function support will be provided, including measures such as mechanical ventilation and continuous blood purification. If the blood pressure is high, providing comprehensive treatment with dilation and diuresis is the main measure while maintaining the water and electrolyte acid–base balance. Once the patient’s condition stabilizes, she is transferred to the obstetrics department for further treatment.

### Implementation of the study

RRI, pCysC, and Scr were measured for all enrolled patients within 12 h of transfer to the ICU, marked as D1. It was repeated every 24 h twice, marked as the second day (D2) and the third day (D3), during which continuous hourly UO monitoring was performed. Hourly UO data were extracted from the ICU information system, and a dedicated person was responsible for recording the data. Because patients in the study were transferred from the obstetrics department to the ICU after pregnancy termination, the cutoff value for initiation of Scr within 48 h could not be determined. Combined with the diagnostic criteria of AKI [5], in this study, PR-AKI was diagnosed considering the change in Scr or UO on D1 after transfer to the ICU. Patients with AKI were assigned to the study group (Group A) and divided into stages I, II, and III, and patients without AKI were assigned to the control group (Group B).

### Statistical analysis

Continuous variables are presented as means and standard deviations or medians and interquartile ranges, and categorical variables are summarized as counts and percentages in each category. Continuous variables were tested using the t-test or Wilcoxon rank-sum test, and categorical variables were tested using the chi-square test or Fisher’s exact test. Univariate and multivariate logistic regression analyses of AKI-related factors were performed. Receiver operating characteristic curve analysis was used to study pCysC and RRI, and the area under the curve (AUC) was compared. Statistical significance was set at P < 0.05. All statistical analyses were performed using SPSS version 23.0.

## Results

During May 2016–June 2021, 182 women admitted to the ICU met the enrollment criteria: 125 cases of pre-eclampsia or eclampsia; 19 cases of placenta previa; 12 cases of placental abruption; 12 cases of hemolysis, elevated liver enzymes, and low platelet count (HELLP) syndrome; 10 cases of postpartum hemorrhage; and 4 cases of amniotic fluid embolism.

Among the 182 patients enrolled in this study, 35 (35/182, 19.2%) were diagnosed with AKI: 23 (23/35, 65.7%), stage I; 9 (9/35, 25.7%), stage II; and 3 (3/35, 8.6%), stage III. Of all AKI patients, six (6/35, 17.1%) had a postpartum hemorrhage and two had AKI stage III. Three AKI stage III patients were excluded from the analysis to avoid any impact on research data because of the requirement of continuous blood purification treatment. Finally, 179 patients were included, 32 in Group A and 147 in Group B.

The Scr, pCysC, and RRI on D1, D2, and D3 in Group A increased, but they tended to decrease gradually over time. Compared with the corresponding values in Group B, there were significant differences in each period (P<0.05) (Table 2).

According to the AKI diagnostic staging criteria, all patients in Group A met the diagnostic criterion of Scr concentration. However, only approximately one-third of the patients met the diagnostic criterion of UO. pCysC and RRI increased in most patients in Group A, especially on D1. There were significant differences in Group A compared with Group B (Table 3).

Through univariate and multivariate logistic regression analyses of AKI-related factors, the three variables of pCysC, RRI, and the combination of pCysC and RRI all independently correlated with AKI. Compared with the value of the indicators alone, the combination of pCysC and RRI increased the predictive value of AKI (OR, 4.026; 95% CI: 10.06–15.46) (Table 4).

A large proportion of the enrolled patients used diuretics during treatment, which greatly interfered with the use of UO to diagnose AKI. Therefore, the RRI and pCysC specificity and sensitivity on D1 were calculated relative to Scr on D1. When serum pCysC was >1.03 mg/L, the AUC for predicting PR-AKI was 0.951 (p<0.001), indicating that pCysC was significant in predicting PR-AKI, with an accuracy rate of 95.1%. When RRI was > 0.64, the AUC for predicting PR-AKI was 0.849 (p<0.001), indicating that the RRI is meaningful in predicting PR-AKI, with an accuracy rate of 84.9%. The AUC of the two combined methods for predicting PR-AKI was 0.979 (p<0.001), indicating that the combination of pCysC and RRI was significant in predicting PR-AKI, with an accuracy rate of 97.9%. These indicators have significant predictive efficacy and high sensitivity and specificity for PR-AKI (Table 5, Figure 1).

**Figure 1.**
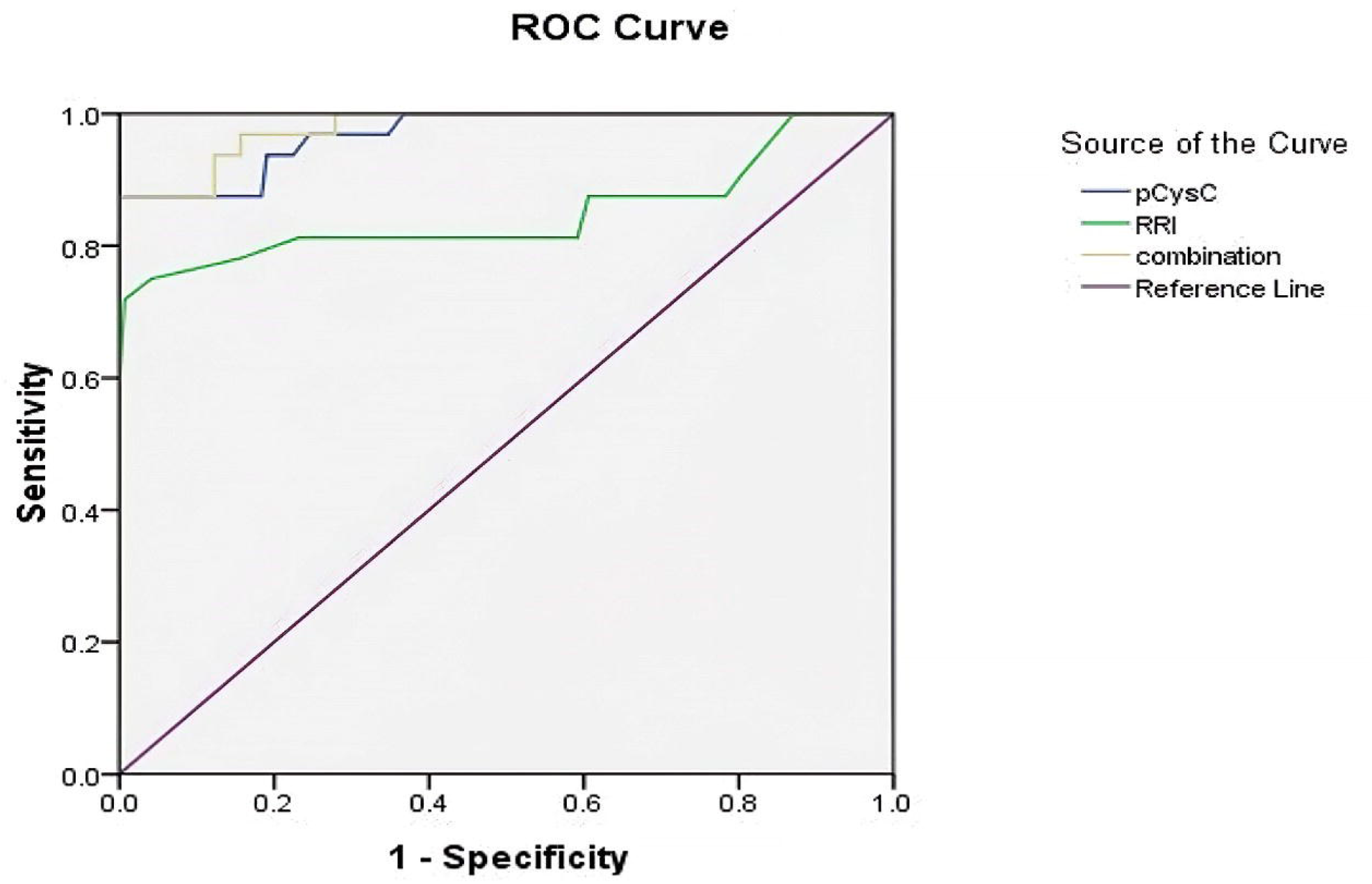
ROC curve of pCysC, RRI, and combined detection in the diagnosis of PR-AKI

The hospital mortality rate of these two groups was determined using Fisher’s exact test, and there was no difference in hospital mortality between the two groups, although the ICU and hospital length of stay in Group A were longer. (Table 6).

## Discussion

The incidence of PR-AKI in this study far exceeded that reported in China [12]; in this study, the patients were enrolled on account of presenting with pathological obstetrics, and the incidence was also related to the severity of the patients’ condition [26-28]. In this study, the APACHE II score was 8 (7–9) in Group B and 10 (8.25–11) in Group A. Some pregnant women with relatively mild conditions were excluded because the ICU stay was < 72 h and most did not have AKI. This may also be related to differences in ICU admission standards in different countries and regions. The purpose of this study was to determine the predictive value of pCysC and the RRI for diagnosing PR-AKI, but the incidence of AKI in pregnant women admitted to both the general ward and ICU in our hospital was not determined.

Mean arterial pressure affects renal perfusion, which affects the RRI results [29]. In the two groups, the mean arterial pressure was higher in patients with eclampsia or pre-eclampsia as the main disease. However, the RRI in the control group was within the normal range, while the RRI in Group A was higher than that in Group B. Moreover, Group A was more affected by postpartum hemorrhage, and the mean arterial pressure was lower than that in Group B. With or without statistical significance between the two groups, other basic indicators were all pathological obstetric-related changes. For example, plasma albumin concentrations were significantly lower in patients with eclampsia or pre-eclampsia. These indicators had no material impact on the indicators in subsequent studies (Table 1).

**TABLE 1.** Comparison of basic clinical and blood biochemical data between group A and group B after ICU admission

| Characteristics | Group B ( n, 147 ) | Group A ( n, 32 ) | t/z | P-value |
| --- | --- | --- | --- | --- |
| Age (yrs) | 30.74±4.85 | 31.59±4.26 | -0.919 | >0.05 |
| BMI ( kg/m<sup>2</sup> ) | 26(25-28) | 26(24-28) | -0.116 | >0.05 |
| MAP ( mmHg ) | 107.57±11.96 | 102.53±15.76 | -2.033 | <0.05 |
| WBC ( ×10<sup>9</sup>/L ) | 13.89±4.54 | 14.37±3.25 | 0.571 | >0.05 |
| Hb ( g/L ) | 133(121-142) | 125.5(83.75-137.5) | -2.655 | <0.05 |
| Alb ( g/L ) | 28.16±4.47 | 26.44±3.39 | -2.056 | <0.05 |
| PH | 7.38(7.37-7.4) | 7.38(7.35-7.39) | -1.349 | >0.05 |
| Blood lactic acid | 1.1(0.8-1.3) | 1.25(0.825-1.8) | -1.945 | >0.05 |
| CRP ( ug/ml ) | 127±42.58 | 69.47±40.45 | 7.222 | <0.001 |
| APACHEII | 8(7-9) | 10(8.25-11) | -5.071 | <0.001 |

Table 2 shows that the Scr concentration in Group A was higher than that in Group B during the first 3 days. With the intervention of ICU measures, Scr concentration showed a gradually decreasing trend and was close to normal on D3. Although the pathogenesis of PR-AKI involves more factors than other causes of AKI [30,31], such patients are relatively young with good basic organ function and strong compensatory ability. After pregnancy termination, the basic cause of PR-AKI was determined, recovery was rapid, and the overall outcome was good. All AKI patients in Group A mainly had AKI stage I. There were only three cases of AKI stage III, two of which were caused by postpartum hemorrhage, which illustrated a feature of PR-AKI from another perspective.

**TABLE 2.** Comparison of Scr (ummol/L), pCysC (mg/L), and RRI between group A and the control group within 3 days of ICU admission

| Groups | D1 |  |  | D2 |  |  | D3 |  |  |
| --- | --- | --- | --- | --- | --- | --- | --- | --- | --- |
|  | Scr | pCysC | RRI | Scr | pCysC | RRI | Scr | pCysC | RRI |
| A | 184.63±27.44 | 1.91(1.82-2.11) | 0.76(0.72-0.78) | 153.25±25.18 | 1.48(1.37-1.66) | 0.7(0.65-0.72) | 110±11.38 | 1.18±0.2 | 0.64(0.61-0.66) |
| B | 93.12±10.78 | 0.83(0.76-0.92) | 0.63(0.6-0.68) | 93.16±8.11 | 0.84(0.78-0.91) | 0.61(0.6-0.65) | 90.64±8.09 | 0.84±0.12 | 0.61(0.6-0.64) |
| t/z | 18.551 | -8.322 | -6.199 | 13.35 | -8.035 | -5.72 | -11.64 | 8.909 | -3.29 |
| P-value | <0.001 | <0.001 | <0.001 | <0.001 | <0.001 | <0.001 | <0.001 | <0.001 | <0.001 |

In Group A, pCysC and RRI, the indicators of renal function monitoring, showed the same characteristics as the changing trend of Scr, which laid the foundation for the clinical application of these two indicators. A previous study showed that an increase in pCysC precedes that in Scr [32]. Because all patients in this group were still critically ill after pregnancy termination and were transferred to the ICU, those in Group A had an increased Scr, which did not reflect the advantage of an earlier increase in pCysC. In pathological obstetrics, the increase in abdominal pressure and blood volume and imbalance of the renin–angiotensin–aldosterone system may increase the RRI values. However, with pregnancy termination, Group B showed a normal range of RRI. In Group A, the RRI increased and showed the same characteristics as the changing trend of pCysC and Scr, which indicated the predictive value of the RRI in AKI. Moreover, the RRI based on ultrasound Doppler is noninvasive point-of-care monitoring, which has stronger dynamics and timeliness and has incomparable advantages over other indicators.

In line with the AKI diagnostic staging criteria, tables 3, 4, 5, and Figure 1 show that although Scr or UO can help diagnose AKI in pathological obstetric patients, especially in critically ill patients with pre-eclampsia or eclampsia as the main research population, the use of diuretic drugs is a common treatment method to reduce hypervolemia and eliminate edema. In this case, it was no longer feasible to diagnose PR-AKI using UO alone. Only 34.4% of those in Group A met the UO standard for the first measurement and evaluation in the ICU. In the current clinical diagnosis and treatment, Scr is the most important and most dependent indicator for PR-AKI diagnosis. However, Scr is not a sensitive indicator. In late pregnancy, the Scr concentration is usually lower than that during the non-pregnancy period. Despite normal Scr concentration, the renal function in late pregnancy may be slightly hampered. Considering this normal concentration could result in a delay in the diagnosis of PR-AKI, which further reduces the sensitivity of Scr in diagnosing PR-AKI. Concerning pCysC and RRI, regardless of the increase in pCysC, RRI, or pCysC and/or RRI, they showed a significant higher increase in Group A than in Group B. These can be used as important supplements in diagnosing PR-AKI, and both have high sensitivity and specificity. Combining these two indicators may be helpful for the timely diagnosis of PR-AKI and early medical intervention.

**TABLE 3.**
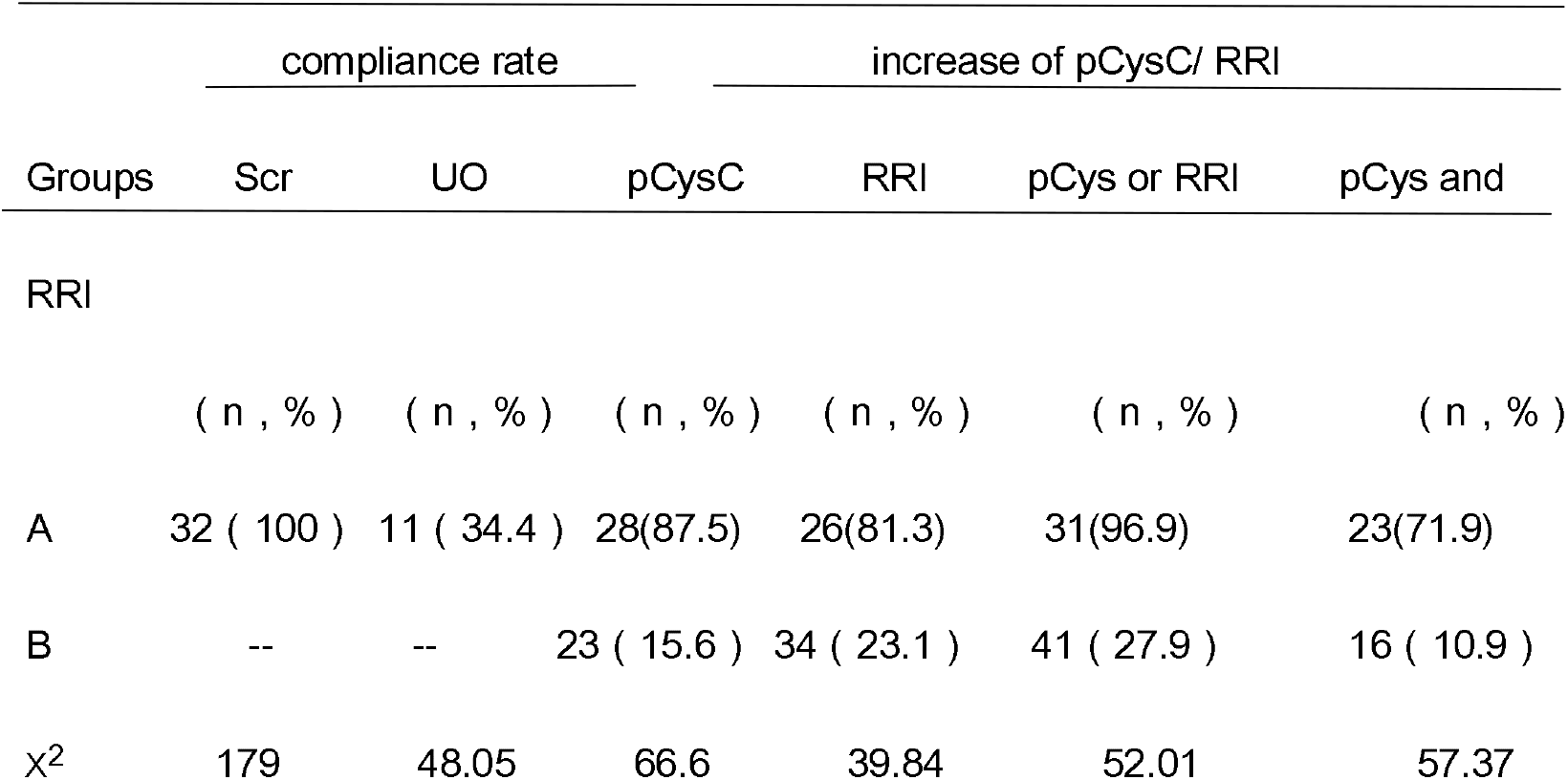
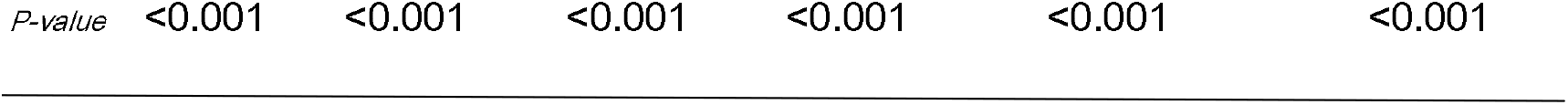
Comparison of the proportion of Scr or UO reaching the diagnostic criteria of AKI and the increase in pCysC/RRI between the two groups according to D1 monitoring results

| Groups | compliance rate |  | increase of pCysC/ RRI |  |  |  |
| --- | --- | --- | --- | --- | --- | --- |
|  | Scr | UO | pCysC | RRI | pCys or RRI | pCys and |
| RRI |  |  |  |  |  |  |
|  | ( n , % ) | ( n , % ) | ( n , % ) | ( n , % ) | ( n , % ) | ( n , % ) |
| A | 32 ( 100 ) | 11 ( 34.4 ) | 28(87.5) | 26(81.3) | 31(96.9) | 23(71.9) |
| B | -- | -- | 23 ( 15.6 ) | 34 ( 23.1 ) | 41 ( 27.9 ) | 16 ( 10.9 ) |
| χ<sup>2</sup> | 179 | 48.05 | 66.6 | 39.84 | 52.01 | 57.37 |

*P-value* <0.001 <0.001 <0.001 <0.001 <0.001 <0.001

**TABLE 4.** Multiple logistic regression analysis of AKI related factors

| Variables | Multivariate analysis |  |
| --- | --- | --- |
|  | Odds ratio (95% CI) | p-value |
| pCysC | 0.002 ( 0,0.018 ) | < 0.05 |
| RRI | 0.003 ( 0,8.932 ) | < 0.05 |
| combination of the two | 4.026 ( 10.06,15.46 ) | < 0.05 |

**TABLE 5.** The sensitivity and specificity of D1 pCysC/RRI in the diagnosis of AKI according to D1 monitoring Scr value

| T pCysC/RRI | sensitivity (%) | specificity (%) | OR-value | 95%CI |
| --- | --- | --- | --- | --- |
| pCysC | 87.50% | 84.35% | 37.739 | 12.091-117.792 |
| RRI | 81.25% | 76.87% | 0.433 | 0.324-0.579 |
| combination of the two | 96.88% | 72.11% | 80.146 | 10.593-606.401 |

Some patients had increased pCysC and RRI but did not meet the current diagnostic criteria for AKI; therefore, they were included in Group B (non-AKI group). The increase in pCysC and RRI and its effects remain unclear for these patients, and a study with a larger sample size is required.

As shown in Table 6, the length of stay in the ICU and the hospital was longer in Group A than in Group B, which was related to the severity of the patients’ condition. Some patients underwent blood purification treatment because of the occurrence of AKI. However, there was no difference in hospital mortality between the two groups because the patients were relatively young and AKI was mainly of stages I and II, which had little effect on the outcome. Whether pCysC and RRI show greater significance in the diagnosis of critically ill patients with poor basic conditions needs to be further studied.

**TABLE 6.** Comparison of clinical outcomes between the two groups

| Outcomes | Group A | Group B | $\chi^2$ or *t*-value | *P*-value |
| --- | --- | --- | --- | --- |
| Length of stay in ICU ( d ) | 5 ( 4.25-6 ) | 4 ( 4-5 ) | 2.031 | <0.05 |
| Length of stay in the hospital ( d ) | 7(6-8) | 6(5-7) | -4.36 | <0.001 |
| Hospital mortality ( % ) | 3.13 ( 1/32 ) | 2.72 ( 4/147 ) | — |  |

This study has some limitations. First, the number of cases in Group A was insufficient, as this was a single-center study. Second, RRI determination had a certain degree of subjectivity. Finally, pregnancy influenced pCysC and RRI. Therefore, further subgroup analysis is necessary with a larger sample size.

## Conclusions

The diagnosis of PR-AKI based only on Scr and UO was insufficient. The RRI and pCysC were important supplements for diagnosing PR-AKI, and the RRI had the advantages of being noninvasive, timely, and dynamic and provide continuous data. The RRI and pCysC had good sensitivity and specificity in predicting PR-AKI, and combining the two was better.

## Data Availability

All data produced in the present study are available upon reasonable request to the authors
All data produced in the present work are contained in the manuscript
All data produced are available online at

## List of Abbreviations

RRI: renal resistance index;
pCysC: plasma cystatin C;
Scr: serum creatinine;
UO: urine output;
APACHE: acute physiology and chronic health evaluation;
AKI: acute kidney injury;
ICU: intensive care unit;
HELLP: hemolysis, elevated liver enzymes, and low platelet count;
POCT: point-of-care testing;
BMI: body mass index;
MAP: mean arterial pressure;
WBC: white blood cell;
Hb: hemoglobin;
AIb: albumin;
CRP: C-reactive protein;
AUC: area under the curve

## Declarations

### Ethics approval and consent to participate

The study was conducted in accordance with the principles of the Declaration of Helsinki, and the study protocol was approved by the ethics committee of Liaocheng People’s Hospital.

### Consent for publication

Not applicable.

### Availability of data and material

With permission from the corresponding author, we can provide participant data and materials for further statistical analysis.

### Authors’ contributions

ST and TW had full access to all data in the study and takes responsibility for the integrity of the data and the accuracy of the data analysis. ST, QW, and ZC contributed to the study design and writing. ZC, MW, ZX, YS, ML, and YC contributed to data acquisition, analysis, and interpretation. HT, XZ, and ML critically revised the manuscript for important intellectual content. HT, XZ, and TW supervised this study. All authors have read and approved the manuscript.

## Acknowledgments

We thank all patients and their families involved in this study. We thank all of those who contributed to this study.

## References

1. Chertow GM, Burdick E, Honour M, et al. Acute kidney injury, mortality, length of stay, and costs in hospitalized patients. [J]. J Am Soc Nephrol, 2005,16(11): 3365–3370. DOI: 10.1681/ASN.2004090740

2. Maced E,Mehta R.L. Preventing Acute Kidney Injury[J]. Crit. Care Clin. 2015, 31(4), 773–784. DOI: 10.1016/j.ccc.2015.06.011

3. Ibrahim A, Ahmed M M, Kedir S, et al. Clinical profile and outcome of patients with acute kidney injury requiring dialysis–An experience from a haemodialysis unit in a developing country[J]. BMC Nephrol. 2016, 17(1), 91.DOI: 10.1186/s12882-016-0313-8

4. Bentata Y, Housni B, Mimouni A, et al. Acute kidney injury related to pregnancy in developing countries: Etiology and risk factors in an intensive care unit[J]. J. Nephrol. 2012, 25(5), 764–775. DOI:10.5301/jn.5000058

5. Levin A, Warnock DG, Mehta RL, Acute Kidney Injury Network Working Group, et al. Improving outcomes from acute kidney injury: report of an initiative[J]. Am J Kidney Dis. 2007 Jul; 50(1): 1–4. DOI: 10.1053/j.ajkd.2007.05.008

6. Barrantes F, Tian J, Vazquez R, et al. Acute kidney injury criteria predict outcomes of critically ill patients[J]. Crit Care Med, 2008, 36(5):1397–403. DOI:10.1097/CCM.0b013e318168fbe0

7. Pickering JW, Endre ZH. The definition and detection of acute kidney injury[J]. J Renal Inj Prev,2013, 30;3(1):21–5. DOI: 10.12861/jrip.2014.08

8. Thomas ME, Blaine C, Dawnay A, et al. The definition of acute kidney injury and its use in practice[J]. Kidney Int, 2015, 87(1):62–73. DOI:10.1038/ki.2014.328

9. Md Ralib A,Pickering JW,Shaw GM, et al. The urine output definition of acute kidney injury is too liberal [J]. Crit Care, 2013, 20;17(3):R112. DOI:10.1186/cc12784

10. Arroyo V. Acute kidney injury (AKI) in cirrhosis: should we change current definition and diagnostic criteria of renal failure in cirrhosis?[J]. JHepatol, 2013,59(3):415–7. DOI:10.1016/j.jhep.2013.05.035

11. Atwal G, Stacey S,Yate P. Ethnicity and acute kidney injury: the correct definition of acute kidney injury?[J]. Br J Anaesth, 2014, 112(1):177. DOI: 10.1093/bja/aet452

12. Liu YM, Bao HD, Jiang ZZ, Huang YJ, et al. Pregnancy-related Acute Kidney Injury and a Review of the Literature in China [J]. Intern Med, 2015;54(14):1695–703. DOI:10.2169/internalmedicine.54.3870

13. Prakash J, Niwas SS, Parekh A, Pandey LK, et al. Acute kidney injury in late pregnancy in developing countries [J]. Ren Fail, 2010, 32(3):309–13. DOI:10.3109/08860221003606265

14. Machado S, Figueiredo N, Borges A, et al. Acute kidney injury in pregnancy: a clinical challenge[J]. J Nephrol. 2012, 25(1):19–30. DOI:10.5301/jn.5000013

15. Song J, Wu W, He Y, et al. Value of the combination of renal resistance index and central venous pressure in the early prediction of sepsis-induced acute kidney injury[J]. J Crit Care.2018 06; 45: 204–208. DOI: 10.1016/j.jcrc.2018.03.016

16. Radermacher J, Ellis S, Haller H. Renal resistance index and progression of renal disease. Hypertension.2002,39(2 Pt 2): 699–703. DOI:10.1161/hy0202.103782

17. Jingchao L, Xiaohua Q, Haibo Q. Application of renal artery resistance index in the assessment of acute kidney injury in critically ill patients. Chin J Intern Med[J], 2014,53(6)496–498.

18. Bagshaw SM, Bellomo, R. Cystatin C in acute kidney injury[J]. Curr Opin Crit Care.2010,16(6): 533–9. DOI: 10.1097/MCC.0b013e32833e8412

19. Maiwall R, Kumar A, Bhardwaj A, et al. Cystatin C predicts acute kidney injury and mortality in cirrhotics: A prospective cohort study[J]. Liver Int, 2018,38 (4): 654 –664. DOI:10.1111/liv.13600

20. Yong Z, Pei X, Zhu B, et al. Predictive value of serum cystatin C for acute kidney injury in adults: a meta-analysis of prospective cohort trials. Sci Rep. 2017, 23, 7: 41012. DOI: 10.1038/srep41012

21. Nejat M,Pickering JW,Walker RJ, et al. Rapid detection of acute kidney injury by plasma cystatin C in the intensive care unit[J]. Nephrol Dial Transplant. 2010, 25 (10):3283–9. DOI:10.1093/ndt/gfq176

22. Soto K,Coelho S,Rodrigues B,Martins H, et al. Cystatin C as a marker of acute kidney injury in the emergency department[J]. Clin J Am Soc Nephrol, 2010, 5 (10):1745–54. DOI:10.2215/CJN.00690110

23. Royakkers AA,Korevaar JC,van Suijlen JD, et al. Serum and urine cystatin C are poor biomarkers for acute kidney injury and renal replacement therapy[J]. Intensive Care Med, 2011, 37(3):493–501. DOI:10.1007/s00134-010-2087-y

24. Obrenovic R, Petrovic D, Majkic-Singh N, et al. Serum cystatin C levels in normal pregnancy[J]. Clin Nephrol, 2011,76(3): 174–9. DOI: 10.5414/cn106792

25. Hamed HM, El-Sherbini SA, Barakat NA,et al. Serum cystatin C is a poor biomarker for diagnosing acute kidney injury in critically-ill children[J]. Indian J Crit Care Med, 2013,17(2): 92–8. DOI:10.4103/0972-5229.114829

26. Lei Y, Nie S, Sun DH, et al. Epidemiology of acute kidney injury in Chinese critical patients[J]. Nan Fang Yi Ke Da Xue Xue Bao, 2016,36(6): 744–50. PMID: 27320872

27. Mas-Font S, Herrera-Gutiérrez ME, Gómez-González C, et al. Epidemiology of contrast-associated acute kidney injury in critical patients. NEFROCON study[J]. Med Intensiva, 2020, 25. DOI: 10.1016/j.medin.2020.07.004

28. Gama RM, Clark K, Bhaduri M, et al. Acute kidney injury e-alerts in pregnancy: rates, recognition and recovery[J]. Nephrol Dial Transplant, 2020, 22. DOI:10.1093/ndt/gfaa217

29. Deruddre S, Cheisson G, Mazoit JX, et al. Renal arterial resistance in septic shock: effects of increasing mean arterial pressure with norepinephrine on the renal resistive index assessed with Doppler ultrasonography[J]. Intensive Care Med, 2007, 33(9): 1557–62. DOI:10.1007/s00134-007-0665-4

30. Taber-Hight E, Shah S. Acute Kidney Injury in Pregnancy[J]. Adv Chronic Kidney Dis, 2020, 27(6): 455–460. DOI:10.1053/j.ackd.2020.06.002

31. Piccoli GB, Zakharova E, Attini R, er al. Acute Kidney Injury in Pregnancy: The Need for Higher Awareness. A Pragmatic Review Focused on What Could Be Improved in the Prevention and Care of Pregnancy-Related AKI, in the Year Dedicated to Women and Kidney Diseases[J]. J Clin Med, 2018, 7(10). DOI:10.3390/jcm7100318

32. Nejat M, Pickering JW, Walker, RJ, et al. Rapid detection of acute kidney injury by plasma cystatin C in the intensive care unit[J]. Nephrol Dial Transplant.2010, 25 (10): 3283–9. DOI: 10.1093/ndt/gfq176

